# Protocol for a qualitative study exploring the experiences and perceptions of dolutegravir/lamivudine dual antiretroviral therapy (the PEDAL Study) in people living with HIV

**DOI:** 10.1101/2021.08.13.21262001

**Authors:** Giovanni Villa, Diego Garcia Rodriguez, David Fray, Amanda Clarke, Caroline Ackley

**Author notes:** **Correspondence to** Dr Giovanni Villa MD MSc PhD, Ground Floor Office, Medical Research Building, Brighton and Sussex Medical School, University of Sussex, Falmer, Brighton, BN1 9PX, E. Authors’ e-mail addresses: Dr Giovanni Villa, Dr Diego Garcia Rodriguez, Mr David Fray, Dr Amanda Clarke, Dr Caroline Ackley.

## Abstract

**INTRODUCTION:** Antiretroviral treatment turned HIV infection into a chronic disease and improved quality of life for people living with HIV. Dual-drug combinations can potentially reduce long-term drug-associated toxicities. We aim to investigate patients’ perceptions and experiences on the safety, effectiveness, tolerability, and unmet needs of the dual-drug combination dolutegravir/lamivudine focusing on patients receiving care in Brighton, United Kingdom. In addition, we will conduct a comparative analysis between patients on dolutegravir/lamivudine and patients on other dual-drug and three-drug combinations. Lastly, the study aims to provide recommendations to improve doctor-patient communication, knowledge and understanding of the treatment plan, and additional care that ought to be considered in patient-centred, holistic care plans.

**METHODS AND ANALYSIS:** Our qualitative methodological framework is based on three main methods: cultural domain analysis, focus group discussions, and in-depth interviews. Cultural domain analysis employs a range of techniques (free listing, pile sorts, and rankings) to elicit terms from informants regarding specific cultural domains (i.e., groups of items that are perceived to be of the same kind). This framework has been co-designed with a patient representative to ensure relevance, suitability, and co-production of knowledge. All methods have been tested to take place online via Zoom®, Skype®, or Microsoft Teams® should participants prefer to participate digitally rather than in person. Padlet®, an application to create online boards, will be used during the cultural domain analysis session. Data collected will be analysed following the completion of each method embracing an iterative approach through applied thematic analysis.

**ETHICS AND DISSEMINATION:** Ethical approval was obtained from the Health Research Authority (Reference 21/NW/0070). Findings will be used to produce recommendations to improve doctor and patient communication by identifying patients’ fears, worries, misconceptions, and general concerns of their drug regimen. Conclusions will be disseminated via journal articles, conference papers, and discussions through public engagement events.

**PROJECT REGISTRATION NUMBER:** IRAS Number: 286277

SPONSOR Number: 076 VIL/ 286277

FUNDER Number: 214249

ClinicalTrials.org Registration Number: NCT04901728

**STRENGHTS AND LIMITATIONS OF THE STUDY:** *Strengths:*

▸ This study will gather qualitative data through three research methods (cultural domain analysis, focus group discussions, and in-depth interviews) to triangulate the findings emerging from patients’ experiences and perceptions of the dolutegravir/lamivudine (DTG/3TC) dual-drug combination
▸ This study is the first of its kind to provide patient-centred insight into DTG/3TC treatment combination to improve clinical care through an in-depth qualitative, iterative, and comparative approach (against previous survey studies on patients’ reported outcomes)
▸ The study’s protocol has been co-designed with a representative of people living with HIV in Brighton and Hove to ensure co-production of knowledge
▸ Data gathered will be analysed through applied thematic analysis to produce recommendations to improve doctor and patient communication after identifying patients’ concerns of their drug regimen
▸ The possibility of taking part in research both in-person and online will allow for increased anonymity and flexibility for patients to participate while simultaneously ensuring that they are safe in the COVID-19 environment by reducing in-person meetings

*Limitations:*

▸ The cohort in Brighton might not be representative of the whole country and groups like women living with HIV, ethnic minorities, and transgender individuals might be underrepresented
▸ Potential participants who might not feel comfortable meeting in person and who lack the digital skills required might be unable to take part in the study
▸ Patients with complex ARV regimens will not be included given their limited treatment options

## INTRODUCTION

Dolutegravir (DTG), an integrase strand transfer inhibitor (InSTI), is currently recommended for both treatment initiation and second/third line therapy for people living with HIV-1 (PLHIV), in combination with either tenofovir disoproxil fumarate/emtricitabine (TDF/FTC), tenofovir alafenamide (TAF/FTC) or abacavir/lamivudine (ABC/3TC).^1 2 3^ The dual-drug combination dolutegravir lamivudine (DTG/3TC) has proven non-inferior to the triple-drug combination TDF/FTC/DTG in the GEMINI-1 and GEMINI-2 trials for the treatment of antiretroviral treatment (ART) naïve individuals.^4^ Treatment guidelines of the European AIDS Clinical Society (EACS), of the US Department of Health and Human Services (DHHS), and of the International Antiviral Society–USA Panel have introduced this option among recommended first-line regimens, providing the patient HIV-1 RNA is <500,000 copies/mL and the CD4 count >200 cells/mm^3^ ^5 6^ Based on the results of the ASPIRE,^7^ LAMIDOL,^8^ and, ultimately, TANGO studies,^9^ the dual-drug combination DTG/3TC has also proven to be a safe and effective option for treatment simplification of ART-experienced suppressed individuals on triple-drug therapy. A recent systematic review and meta-analysis explored real-world effectiveness and tolerability of DTG/3TC in virologically suppressed patients and documented long-term virological outcomes consistent with findings from randomised clinical trials.^10^

Although there is clinical evidence of the safety, effectiveness, and tolerability of dual-drug regimens ^11 12 13 14 15^, there is limited insight into patient experiences and perceptions of dual-drug combinations, including the DTG/3TC regimen. A qualitative study conducted in the United States and in Spain explored patients’ perspectives and experiences in 39 patients on dual-drug combinations and documented that participants viewed dual-drug regimens as a significant and positive advance, in terms of its effectiveness, with reduced toxicity and essentially no reported side effects^16^. The study highlighted the central role of health-care providers in the decision to switch to dual-drug combinations.

The PEDAL study aims at exploring patients’ experiences and perceptions of the dual-drug regimen DTG/3TC, including potentially unmet treatment needs and reported outcomes for those already on this combination. In addition, we will conduct a three-phase comparative study with a control population alongside the target population (Figure 1). The control population will include people living with HIV on dual-drug regimens other than DTG/3TC and a group on triple-drug therapy. In the control group of participants receiving dual-drug therapies, we will include patients (a) on DTG/rilpivirine (RPV), (b) on boosted-darunavir plus lamivudine (DRV/ritonavir[r] or DRV/cobicistat[c] + 3TC), and (c) on boosted-darunavir plus raltegravir (DRV/r or DRV/c + RAL). The addition of the control group of patients on other dual-drug therapies will allow us to tease out the particular characteristics of DTG/3TC beyond the mere reduction of molecules employed for the treatment. This study will be the first of its kind to provide patient-centred insight into this specific treatment combination and to produce recommendations for improved clinical care.

**Figure 1.**
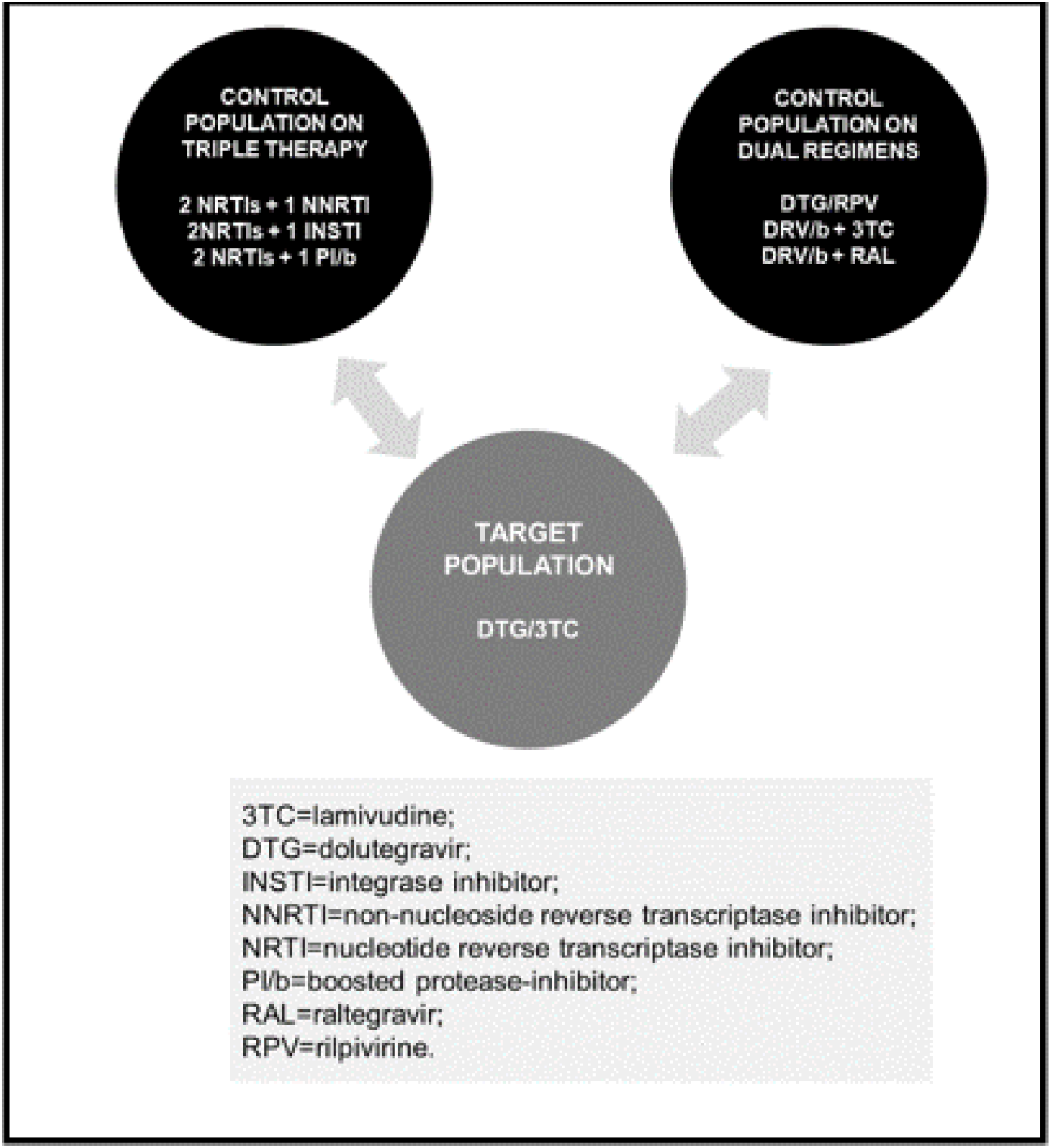
Control populations and target population.

### Aims and objectives

This research will use a qualitative methodology to explore patients’ perceptions and experiences of the dual-drug treatment regimen DTG/3TC, including potentially unmet treatment needs.

The objectives are:

- To investigate patients’ perceptions and experiences of the safety, effectiveness, tolerability, and unmet needs of DTG/3TC;
- To conduct a comparative analysis of the safety, effectiveness, tolerability, and unmet needs between patients on DTG/3TC and patients on other dual-drug and three-drug combinations;
- To provide recommendations to improve doctor-patient communication, knowledge and understanding of treatment plans, and additional care to be considered in patient-centred, holistic care plans

### Research questions

- What are patients’ experiences and perceptions of the safety, effectiveness, and tolerability of DTG/3TC?
- What are the unmet needs of patients taking DTG/3TC?

### Theoretical approach

The theoretical framework that informs this study is rooted in Critical Medical Anthropology (CMA). Central to the CMA approach to the study of health and disease is (1) close attention to ‘ecological, biological and cultural factors;’ (2) consideration of the ‘political and economic forces that influence disease patterns and affect access to health care resources;’ and (3) the ‘opportunity for health-promoting interventions’.^17^ Through this approach we can better understand human psychobiological systems, patients’ experiential responses to illness, their support networks, and physician-patient interactions in relation to the DTG/3TC treatment regimen and future direction of dual-drug HIV treatment.

## METHODS AND ANALYSIS

### Study design

The PEDAL study is a qualitative study containing in-built elements for continuous data analysis based on an iterative approach. This will allow to continuously refine our methodological framework following the implementation of each method. Data collection for this study commenced in June 2021 and is expected to be finalised by April 2022. We will engage with a target population (patients on DTG/3TC) and two control populations (patients on other dual-drug regimens and triple-drug ART) through cultural domain analysis (CDA), focus group discussions (FGDs), and in-depth interviews (IDIs). Upon completion of data collection, applied thematic analysis (ATA) will be used to identify emerging themes.

### Recruitment

#### Sampling technique

The PEDAL study includes a target and control groups. The variables that this study sets out to explore (i.e., safety, effectiveness, tolerability, and patients’ unmet needs) will be better comprehended by including a control group to allow for comparison. However, and in light of our iterative approach, the inclusion of control groups will be evaluated as the study progresses. For example, if the CDA data reveals unique findings for the two and three drug regimens, then we will continue the study with a control group consisting of people on both two and three drug regimens. If the data does not reveal unique findings within the dual-drug regimen control group, then the control population for the FGDs and IDIs will be modified to only include patients on three-drug regimens excluding those on alternative dual-drug regimens.

We will use purposive sampling^11^ to recruit a diverse population in terms of age, gender, socioeconomic status, ART, treatment duration, and number of previous ART regimes. Particular attention will be placed on recruiting participants from underrepresented groups (e.g., age ≥70 years; women; people with history of challenges to adherence; people with co-morbidities). In the event of an insufficient sample recruited via a purposive sampling strategy, we will use snowball sampling^11^ where participants recruit future subjects from their networks (providing they meet the inclusion criteria).

Participants will be recruited through various methods. Firstly, the research team will screen the clinical database to identify individuals to be recruited during clinical sessions. Clinical research nurses will support participant identification and screening to check patient eligibility. They will provide patients with a study information sheet and a researcher will approach the patients to obtain verbal consent should they want to learn more. Additionally, participants will be recruited through peer support groups, women’s groups, online groups, flyers emailed and handed directly to patients by doctors, and to be circulated amongst consultants, nurses, and by the Research Assistant in the clinic. Patients interested in taking part in the study will be offered to discuss the project further with a team member acting as a representative of people living with HIV. We expect that being able to speak with someone they can identify with will reduce hesitation to participate, build confidence, and increase motivation to stay engaged.

Participation will be in-person or online depending on the COVID-19 guidance and participants’ preferences. Participants will be offered up to £30 in vouchers to compensate for their time. They will receive £10 for participation in one method; £20 for participation in two methods; and £30 for participation in all three methods. Additionally, those based in Brighton and Hove will be reimbursed for travel costs.

#### Sample size

Out of a total HIV cohort composed by more than 2,000 patients at BSUH, there are currently 87 patients on the dual drug regimen DTG/3TC. We will approach all 87 patients to participate in the study; however, based on previous study recruitment at our centre we anticipate about 20% of those approached to consent. During the CDA phase, we will recruit a minimum of 8 and up to 40 participants from the target population and 80-116 participants from the control population for a sample range of 80-120 participants. Upon completion of the CDA phase, we will conduct 1-2 FGDs with each group, with each consisting of 6-10 people. Lastly, we aim to conduct 6-12 IDIs with patients from each group.

The sample size used in qualitative research methods is often smaller than in quantitative research. This is because qualitative research is mostly concerned with gaining an in-depth understanding of a phenomenon and is focused on meaning. In-depth interviewing is not necessarily concerned with making generalisations to a larger population and do not tend to rely on hypothesis testing but is rather a more inductive process. As such, the aim of FGD and IDI data is to create analytical, demographic, and ethnographic categories to analyse relationships between categories while attending to the lived experience of participants.^18^ ^19^

We will follow the principle of data saturation, when the data collection process no longer offers any new or relevant data. Conducting interviews, scholars have found that data saturation often occurs within the first twelve interviews while meta-themes might appear after six interviews.^20^ If we determine that saturation has not been achieved in any method, then the time period for recruitment will be extended.

#### Inclusion and exclusion criteria

Adult people living with HIV (PLHIV) (age ≥18 years) will be invited to participate if they have capacity to consent, receive HIV care at the HIV Department of the University Hospitals Sussex, and are on one of the following therapies:

1. **Target population:** DTG/3TC
2. **Control population**
  a. ***Groups on dual-drug therapies:***
    1. DTG/RPV
    2. DRV/r or DRV/c + 3TC
    3. DRV/r or DRV/c + RAL
  b. ***Groups on triple-drug regimens:***
    1. 2 nucleos(t)ide reverse transcriptase inhibitors (NRTIs) + 1 INSTI;
    2. 2 NRTIs + 1 non-nucleoside reverse transcriptase inhibitor (NNRTI);
    3. 2 NRTIs + boosted protease inhibitor (PI/b)

Patients taking DTG/3TC off-label (i.e., with a history of virological failure or suspected resistance to 3TC or DTG) will be excluded. Patients not fitting the inclusion criteria including those taking off-label drug combinations due to complex HIV resistance patterns, ART naïve, or individuals declining ART will be excluded. Patients without access to the technology to take part online will be given the option to participate in person (depending on COVID-19 guidance). If this is not possible, they will be excluded.

### Data collection

We will employ three main methods: CDA, FGDs, and IDIs. Participants will be given the option to take part online or in person. Online participants will be able to join via Microsoft Teams®, Zoom®, or Skype® from a location of their choosing. In-person participants will attend a quiet room at the Clinical Research Facility at the University Hospitals Sussex or at The Sussex Beacon. During FGDs, those attending physically will be informed that everything discussed within the room should remain confidential and that identities cannot be disclosed outside.

#### Cultural domain analysis

CDA is an approach derived from cognitive anthropology to describe the contents, structure, and distribution of knowledge in organised spheres of experience, or cultural domains.^21^ The goal is to understand how people in different cultures (or subcultures) interpret the content of domains differently.^22^ Asking participants to discuss positive and negative factors related to their treatment, cultural domain analysis will focus specifically on exploring the *unmet needs* of patients on dual-drug or triple-drug therapy.

When facilitating CDA sessions, we will employ three tools: free listing, pilesorts, and rankings. Free listing is a simple method where participants are asked to list all they know about a particular topic to get them to mention as many items as they can in a domain. After completing the free listing exercise, we will introduce the pilesort task to elicit judgements of similarity among the items shared during the free listing question.**Error! Bookmark not defined**. The final section will ask participants to rank order the positive/negative factors that most/least meet their treatment needs and support their quality of life while undergoing treatment.

When CDA is conducted in-person we will ask participants to write their answers on notecards and to use these cards to complete the pilesort and ranking tasks. When conducted online, we will utilise Padlet®, a real-time participatory online platform where users can share and organise content to virtual boards called “padlets”. At the start of each session, participants will receive a unique and confidential link to a new Padlet® page. The page will be linked to the researchers’ professional account and no personal data will be linked to the participant. Both the researcher and the participant will log into the same Padlet® page at the same time. This will allow the researchers to observe the participant list, sort, and rank their domains in real time. At the end of the session, the researcher will export the Padlet® board as an image and save it in a secure, password-protected location for analysis.

#### Focus group discussions

After completing the CDA sessions, we will conduct data analysis in line with our iterative approach. We will then hold 1-2 FGDs with each group, in-person or online. In the FGDs with patients on DTG/3TC we will elicit experiences on the treatment and perceptions before switching to it. In the FGDs with patients on other dual and triple-drug therapy we will explore perceptions of DTG/3TC, of ART, and of the future of HIV care and treatment generally.

Each FGD will have a moderator and a facilitator. Due to potential hesitancy to disclose their status to others, online participants will be able to choose if they want to have their video on or off and if they would like to use a pseudonym.

#### In-depth interviews

Following completion of the FGDs, the research team will analyse the data to refine interview questions. After this, we will conduct 6-12 IDIs with participants on DTG/3TC to elicit narratives and treatment histories of the regimen. IDIs with the target population will allow participants to share specific experiences, fears, hopes, concerns, and unexpected outcomes of the therapy. Interview data will be analysed according to the variables of *safety, tolerability*, and *effectiveness* to provide case studies of patients’ experiences. We will also conduct 6-12 IDIs with participants on alternative dual therapy treatments, and 6-12 IDIs with participants on triple-drug therapy regimens. This will allow us to learn about potential misconceptions, misunderstandings, rumours, and knowledge gaps around DTG/3TC from patients on alternative therapies.

### Data analysis

Quantitative data emerging through CDA will be analysed through ANTHROPAC®, a software used to collect and analyse data on cultural domains. We will conduct proximity analysis to compute measures of similarity and difference between respondents on DTG/3TC against dual-drug therapy and triple-drug therapy. Audio-recordings from FGDs and interviews will be transcribed to be coded on NVivo.

Data will be analysed using ATA to identify implicit and explicit ideas. Defined as “a method for identifying, analysing and reporting patterns (themes) within data”^23^, it has been used in public health research to address issues that are practical or applied in nature.^24^ In ATA, the researcher identifies key themes that are transformed into codes. Following an analysis of each group, a second stage analysis will be conducted to compare and contrast findings across groups. The analysis will seek out consensus, disagreement and inconsistency among participants.

## ETHICS AND DISSEMINATION

### Ethical considerations

When participants are identified by members of the healthcare team, they will seek oral consent to contact them with further information about the study. Once participants have orally agreed to participate in an eligibility screening or for us to share further information about the study, we will begin the process of obtaining informed consent. We will give the participants an information sheet and consent form, which they will be asked to return signed and dated before proceeding to the next phase. Prior to any CDA, FGDs, and IDIs the researcher will again seek oral consent to ensure the participant has read and understood the information sheet and consent form.

Participants will be invited to join all components of the research. However, they may choose to take part only in the first method, the first and second method, or all three methods. If participants take part in more than one method, both informed written and oral consent will be taken prior to participating in each of the sessions. Ethical approval has been obtained from the HRA Research Ethics Committee (REC reference: 21/NW/0070).

### Data protection and patient confidentiality

All investigators will comply with the requirements of the Data Protection Act (2018) with regards to collection, storage, processing and disclosure of personal information and will uphold the Act’s core principles. Participants’ information will be replaced by an unrelated unique sequence of characters to guarantee anonymisation. A reconciliation list will be created and stored in a locked cabinet with the study protocol at the Clinical Research Facility at the University Hospitals Sussex, whereas anonymised data collected via the audio-recording of the FGDs and the IDIs, and the images from the CDA will be saved in a folder on the University of Brighton OneDrive, which only the research team will have access to.

## OUTPUT AND DISSEMINATION

Study findings will be disseminated via journal articles, conference papers, and discussions through public engagement events with the Brighton and Sussex Medical School and the Sussex Beacon including recommendations to be used in practice. These activities will contribute to informing the future of HIV treatment by providing evidence of patients’ perceptions and experiences of dual-drug regimens. Ultimately, we expect to improve doctor and patient communication by identifying patient fears, worries, misconceptions, and general concerns of their drug regimen.

## DISCUSSION

Despite the existence of clinical evidence of the safety, effectiveness, and tolerability of dual-drug regimens**Error! Bookmark not defined**., there is currently limited insight into patient experiences and perceptions of dual-drug combinations, including DTG/3TC. Our study will therefore contribute to knowledge by exploring patients’ experiences and perceptions of dual-drug regimens, including potentially unmet treatment needs and reported outcomes for those on this drug combination.

### Limitations

The selected target and control populations involve a potential limitation because of the lack of engagement with other groups excluded from the study (i.e., those taking off-label drug combinations due to complex HIV resistance patterns, ART naïve, or declining ART). Moreover, the Brighton cohort is largely composed of men who have sex with men, hence might not be fully representative of other contexts in the UK. All efforts will be in place to recruit under-represented groups: for example, to ensure an adequate enrolment of women, we will target a dedicated service for women living with HIV at the Lawson Unit (the Sunflower Clinic). No formal matching of patients between groups will take place, and this might generate unbalances that could potentially influence the findings. All efforts will be in place to balance the groups according to age, gender, and ethnic background to mitigate this risk.

The current COVID-19 pandemic has been limiting face-to-face interactions in routine clinical practice in the attempt to contain the spread of the virus. We have overcome this barrier by designing online digital methods, however potential participants who do not have access to the required technology to participate and that cannot or do not wish to take part in person will be excluded. Despite the potential limitations of digital methods for less tech-savvy participants, this approach presents benefits such as increased anonymity due to the option to turn one’s camera off or use pseudonyms. Additionally, accessing the online venue might be less of a barrier to participation than finding time to travel to the research location.^25^

## CONCLUSIONS

The PEDAL study presents a unique opportunity to identify patients’ perceptions and experiences of the dual-drug regimen DTG/3TC, including potentially unmet treatment needs. Our qualitative methodological framework including the three key methods of CDA, FGDs, and IDIs has been designed to take place either physically or online considering the current COVID-19 context. This will contribute to producing knowledge on digital research while identifying the needs of patients to produce recommendations that improve doctor-patient communication, knowledge and understanding of treatment plans, and additional care that ought to be considered in patient-centred care plans.

## Data Availability

The manuscript is a protocol paper and as such does not present data.

## AUTHORS’ CONTRIBUTIONS

GV is the Chief and Principal Investigator of the study, he conceptualised the study and wrote the protocol with CA; DGR is a Research Assistant, he converted the protocol into its publishable format and co-authored the protocol; DF and AC are Co-Investigators and co-authored the protocol; CA is the co-Principal Investigator of the study, she conceptualised the study and wrote the protocol with GV. All authors supported the development and critical review of the protocol and of this manuscript.

